# Problematic gaming in occasional, low, intermediate or heavy use gamers – not all screen time is created equal – a reanalysis

**DOI:** 10.64898/2026.07.30.26359383

**Authors:** Benoit Bediou, Sezen Cekic, Douglas A. Gentile, Daphne Bavelier

**Affiliations:** Université de Genève, FPSE, 40 boulevard pont d’Arve, 1205 Genève, Switzerland; Fondation campus biotech, 9 chemin des mines, 1211 Genève, Switzerland; Iowa State University, 901 Stange Ave, Ames, IA USA; Digital Wellness Lab, Boston Children’s Hospital

**Keywords:** problematic gaming, longitudinal risk and protective factors, media consumption, impulsivity, specification curve analysis, linear mixed effects model

## Abstract

Despite having received increasing attention over the past decade, little is known about the factors that can increase or decrease symptoms of problematic gaming (PG) in individuals with different video gaming (VG) habits (e.g., occasional use gamers vs heavy use gamers). Here, we apply a specification curve analysis to assess the impact of changes in two key risk factors, gaming consumption and impulsivity, as well as one protective factor, family environment, on changes in PG symptoms over 2 years in a large sample of Singaporean adolescents between 2007 and 2009. By considering individual differences in baseline video game habits, we show that changes in VG consumption influenced changes in PG in adolescents who reported not usually playing games at study inception (i.e., occasional use gamers), whereas changes in impulsivity influenced PG in those who reported being the most heavy usegamers at study inception. These findings highlight the importance of designing preventative measures that differ as a function of current video game habits. Possible measures may involve introducing gaming progressively (e.g., through setting age-appropriate limits or structured scaffolding with parents) to improve tolerance and reduce preoccupation in occasional gamers while promoting self-control early in development before gaming can become a problematic habit.

## Introduction

Since it was first introduced in the research appendix of the DSM-V [1] as a condition for further study, Internet Gaming Disorder (IGD) has garnered increasing attention in adolescent mental health due to its potential impact on psychological well-being and social functioning [2]. We refer here to problematic gaming (PG) as signaling problematic behaviors around video game use. PG pre-dates IGD (following the DSM-5/ APA 2013 terminology) or ‘Gaming Disorder’ (GD) according to the 11th Revision of the International Classification of Diseases (ICD-11, WHO, 2022) [3,4]. While PG and IGD rely on a similar screening tool (with 9 of the10 PG scale questions being shared with the IGD scale), PG only aims at capturing the severity of self-reported symptoms around gaming, making it amenable to clinical and sub-clinical populations, when IGD refers to a clinical diagnosis. Accordingy, the focus of this work on that broader construct of PG and relies on self-reported PG rather than clinical diagnosis, as is common in the field. Studies to-date, despite highlighting variations in prevalence of PG diagnosis [5], point to links between PG and a number of underlying factors contributing to its development and persistence. Research into its causes often relies on the epidemiological triad model, which posits that health problems arise from the interaction among the agent (game), the host (individual), and the environment (social/familial context). In the context of video game consumption, the agent involves specific game design characteristics, such as mandatory social interactions or multiplicity and variable schedules of rewards [6], which can foster maladaptive player-game relationships [7,8]. The host encompasses individual and intrapersonal factors like depression, anxiety, attention problems, or impulsivity (e.g., [9,10]). The environment includes interindividual aspects like parent–child relationships and social support (e.g., [11,12]). Investigating these complex interactions is crucial not only for our understanding of how PG arises but also for developing effective prevention and treatment strategies.

### Modifiable risk and protective factors of PG

Although PG is inherently multivariate, recent studies, including systematic reviews, and meta-analyses focusing on longitudinal effects have consistently identified several core, modifiable factors that warrant attention, as they represent ideal targets for intervention [13,14]. Commonly cited are playing time, impulse regulation, psychopathological and cognitive factors and interpersonal factors. We review each in turn.

Excessive playing time is one of the most consistently examined intrapersonal predictors. Heavy gaming, defined in some longitudinal cohorts as playing > 240 minutes (4 hours) per day during weekdays, is a significant longitudinal risk factor, demonstrating high adjusted rate ratios for both the incidence (aRR = 2.03) and persistence (aRR = 2.63) of PG. Such extreme usage may be interpreted as a core feature of PG, specifically the “loss of control” feature of PG [15]. The pooled correlation for gaming time associated with subsequent symptom severity is strong (r = 0.31) in longitudinal studies [13], according to the benchmarks for interpreting longitudinal (cross-lagged) effects (0.03, 0.07 and 0.12 respectively for small, medium and large effects) [16]. Other game-related risk factors include a preference for multiplayer games and for online games [15], including some debates regarding which game genres may show greater association with PG [17–20].

Secondly, constructs related to executive control and self-regulation are also critically important. High self-control acts as a strong longitudinal protective factor against PG (r = -0.27), while high impulsivity is a consistent risk factor (r = 0.11) in longitudinal cohorts [13]. These concepts reflect difficulties in behavioral inhibition associated with the inability to resist the urge to game and may differentiate between disordered versus non-disordered, while still heavily gaming [21]. Here, we refer to impulsivity as a multidimensional construct which encompasses both positive and negative facets of impulsive behavior, perseverance, and sensation seeking. The five dimensions of impulsivity are (1 & 2) negative and positive urgency (acting rashly when experiencing intense negative or positive emotions respectively, and thus exhibiting poor self-control), (3) lack of premeditation (acting without considering the consequences, or poor pro-active control again), (4) lack of perseverance (failing to maintaining focus on a boring or difficult task), and (5) sensation seeking (pursuing exciting activities). In this framework (known as the UPPS-P model, [22]), impulsivity reflects a tendency to act precipitously, to overlook alternative courses of action, and to disregard consequences, and it is a recognized predictor of maladaptive behaviors that arise from poor regulation of positive and negative emotions (see [23]).

When conceptualizing the effects of impulsivity and lack of self-control on PG, the I-PACE model is also worth considering [24]. According to the I-PACE model, impulsivity may also contribute to other PG risk factors. For example, both ruminations about gaming and gaming to escape reality (i.e. as a coping mechanism) may signal high impulsivity or low emotion / impulse regulation abilities. Consistent with the idea that impulsivity and self-control are two sides of the same coin, Warburton et al. 2022 reports an odds ratio of OR = 1.12 for impulsivity and 0.87 for self-control [25], and the meta-analysis by Ji et al. 2023 [26] reports r = 0.32 for impulsivity (risk factor), and r = - 0.31 for self-control (protective factor). A positive association between increased impulsivity and PG was reported by 32/33 studies [27], and associated with a pooled effect-size of r = 0.29 [28], which is small-to-moderate for cross-sectional studies.

A third risk factor for PG concerns pre-existing psychopathology, such as ADHD (pooled OR = 1.4) [29]. ADHD is also a significant predictor of the persistence of high risk PG over a 2-year period (aRR = 2.14) [15]. Additionally, maladaptive cognitions (e.g., short-term thinking, rumination, avatar attachment), gaming motivations (e.g., gaming to escape reality, to experience flow or to cope with negative emotions), and dysregulated behaviors (e.g., rule-breaking, deceiving) are also strongly correlated with PG symptoms [30]. As discussed in the I-PACE model, each of these behaviors can be related to impulsivity / lack of self-control [26].

A fourth dimension, beyond agent (game-based factors) and host (individual traits), influencing PG is interpersonal factors. Interpersonal factors range from the immediate family environment to peers at school and to more remote social groups (e.g., mentors, teachers, or more distant peers). Interpersonal factors play a key moderating role in PG [11]. For example, parental withdrawal (lack of emotional support) correlates positively with PG symptomatology (r = 0.28), functioning as a risk factor [12]. Conversely, strong parent–child relationships and high social support are protective, showing negative associations with PG in longitudinal studies (social support: r = -0.14; parent-child relationships r = -0.15; parental supervision: r = -0.09) [13]. Lower attachment to parents and less open communication are also associated with higher risk for PG. Cross-sectionally, factors such as family closeness (r = -0.19, [14]) and family connectedness correlate negatively with PG symptomatology (standardized beta = -0.16), with the latter being associated with lower odds of PG (OR = 0.50) [25].

### The Critical Gap: Heterogeneity and the Non-Linearity Hypothesis

Despite the consistent identification of gaming time as a central variable, prior cross-sectional studies exhibit substantial heterogeneity in reporting the magnitude of the correlation between weekly video game consumption and PG symptoms. Odgers and Jensen [31] reviewed the extent of evidence linking video game play with PG in (a) systematic reviews and meta-analyses, (b) large-scale preregistered cohort studies and (c) intensive longitudinal and ecological momentary assessment studies. They concluded that evidence was scarce and of questionable practical significance, because the associations (i) were generally small, and (ii) did not distinsguish between cause and effect. For instance, the association between video game consumption and PG was very low (e.g., r = 0.03) in a cross-sectional French sample of N = 418 young adults [32], whereas another large-scale study involving 123,262 gamers from 168 countries (recruited in the context of the smart gaming campaign) documented a much stronger dose-response relationship, with increases in weekly time spent gaming accounting for 10% of the variability in PG symptomatology [33]. Across studies, the association between gaming time and PG is not systematic, highly variable, and generally weaker once adjusted for confounds such as age, sex, educational level, SES, geographical origin, and type of population studied [34–37].

Such heterogeneity may result from assuming a simple linear relation between time spent gaming and negative outcomes. Recent theoretical and empirical work suggests a possible curvilinear relationship [38]. This perspective, sometimes referred to as the “Goldilocks hypothesis,” suggests that psychological and cognitive outcomes may be optimal at intermediate levels of digital engagement, while both non-users and extremely heavy users may experience detriment [38,39]. If true, this non-linearity could explain why studies sampling populations with predominantly low-to-intermediate gaming hours (e.g., general population cohorts) find low correlations, while studies focusing on highly engaged gamers find much stronger correlations. Additionally, studies that have focused on a single video game community have generally reported small correlations between gaming time and PG [40–42].

Several studies have also focused on gamers across gaming genres including from Massive Multiplayer Online Role Playing Games (MMORPG) or Multiplayer Online Battle Arena games (MOBA). Hussain & Griffith [42] found a positive correlation between excessive online gaming (which they defined as 35 hours or more of game play per week) of MMORPGs and psychological and behavioural ‘dependence’ measured with an adapted addiction scale (*r* = .14, *p* < .05). Achab et al. [40] found that problematic players of MMORPGs reported a higher number of hours spent on the internet compared to non-problematic players of MMORPGs; however, this effect had a negligible effect size on PG (OR = 1.18 and 1.28, corresponding to r = .05 and .07).

A non-linear relationship between gaming time and PG could explain, at least in part, the large variability across cross-sectional studies linking PG to hours of game played, which has been reported to vary between values as low as r = 0.03 - N=418 [43] and as high as r = 0.77 - N=1,003 [44]. Although both studies recruited online gamers, the sample characteristics differed. The former, Laconi et al. [43] included 51% men with mean age of 21.9 and an averge weekly gaming time of 14.8 hours (*SD* = 17.2), whereas the gamers in the latter, Pontes et al. [44], included 85.2% males with a mean age of 26 years, and a mean gaming time of 23.24 (SD = 16.55) hours per week. These studies may have sampled different populations corresponding to different portions of the same non-linear relationship, resulting in different linear estimates.

To date, few studies, particularly longitudinal ones, have accounted for how a player’s initial habitual gaming level might moderate the impact of established risk and protective factors on the subsequent progression of PG. Leveraging a longitudinal design allows for a critical test of this moderation hypothesis by establishing the temporal precedence of predictors [29].

### Study aims and novelty

Here, we use a longitudinal dataset [45] to quantify the impact of naturally occurring changes in video game consumption (VG) and other key modifiable risk and protective factors (impulsiveness and family environment) on the onset or progression of PG symptoms over two years. Our methodology allows for the investigation of temporal associations while acknowledging the continuous nature of PG symptoms. The aims of the present study are two-fold. First, we examine the relations between prospective changes in VG consumption, impulsiveness, and family environment and subsequent changes in the severity of self-reported PG symptoms. Second, we test the hypothesis that the strength of these associations is significantly moderated by initial gaming habits, assessed at baseline, by categorizing participants into distinct groups (occasional, low, intermediate, or heavy gaming).

We hypothesize that this heterogeneity stems from the failure to account for an underlying non-linear relationship and individual differences in initial gaming habits. More specifically, we hypothesize that (1) increases in VG consumption and impulsiveness will be positively associated with increases in PG symptoms, while improvements in the family environment will be protective; and (2) the predictive strength of these factors will vary significantly depending on the baseline gaming habit group, consistent with the hypothesized non-linear model [38]. Crucially, we do this by modeling individual differences in video game consumption (occasional, low, intermediate, or heavy gaming) at study inception, treating the level of habitual use as a central moderator of risk. Our approach provides a powerful framework for distinguishing risk factors across different player profiles. Critically, we distinguish between adolescents who, at study inception, report not usually playing video games at baseline (occasional gaming; i.e., they answered “no” when asked whether they usually play), from those reporting between 1 and 10 hours/week (low gaming), between 10 and 28 hours/week (intermediate gaming), or more than 28 hours/week (heavy gaming), closely corresponding to the suggested APA cutoff of 30 hours/week for problematic gaming [1]. Throughout, these four categories are used as engagement-based group labels — a shorthand for adolescents grouped by their baseline weekly play — rather than clinical categories or fixed identities, and this convention applies equally to all figures and tables, including those in the supplementary information. By doing so we can begin to unravel the game play habits contributing to the development and maintenance of PG, crucial information if one is to devise prevention strategies based on media consumption [45].

## Materials and Methods

We reanalysed a dataset used in several publications. A full description of the data collection procedures is available in Gentile et al. 2011 [47]. For this study, data from 3,434 students from 6 primary and 6 secondary schools in Singapore were used. Participants completed a survey comprising 10 topics covering various areas including, socio-demographic information, PG, gaming habits, personal strengths, social attitudes, aggression and hostile traits, gaming experiences, home environment and parental control, and somatic symptoms. Anonymous data for this reanalysis were accessed on March 6^th^, 2023. The authors didn’t have access to information that could identify participants. The questionnaires were administered as paper-and-pencil surveys in participants’ classrooms by their teachers, who followed detailed standardized instructions; four counterbalanced questionnaire versions were distributed across classes. Schools constituted a convenience sample: principals of twelve schools (six primary and six secondary) accepted an invitation to participate, and the participating classes were selected by each school. Participation was voluntary, students could withdraw at any time, and no reimbursement was provided.

### Dataset

Most of the cohort was followed over a three-year period with three data collection waves spaced by one year. All students within the general education program at the schools were eligible to participate, and approximately 99% of eligible students participated. Data were collected from 2007 to 2010 (i.e., 1-year lag between waves).

### Measures included in this reanalysis

The measures included in this reanalysis are listed below. The complete list is available in Choo et al. 2010.

#### Problematic gaming

Participants answered a screening instrument adapted from the pathological-gambling criteria of the DSM-IV. The 10-item DSM-IV-derived composite for PG has been validated and used in multiple peer-reviewed analyses of this and related cohorts [52–55]. The DSM-V IGD criteria (APA, 2013) were themselves developed by adapting nine of the ten DSM-IV pathological-gambling criteria to gaming [50]; accordingly, nine of our ten items map onto the DSM-V IGD criteria (Table S6). The remaining item (co-dependence — needing to borrow money to game) is the one DSM-IV PG item without a DSM-V IGD analogue. Participants could respond “no,” “sometimes,” or “yes” to each of the 10 symptoms and we summed all answers coding yes as 1, no as 0 and sometimes as 0.5. The dependent variable is the sum of all 10 items which are available in Table S2 and represents a measure of the severity of PG symptomatology. Briefly, the ten items index the core symptoms of the construct — preoccupation, tolerance, withdrawal, loss of control (unsuccessful attempts to cut back), loss of interest in other activities, continuation despite problems, deception about gaming, escape/mood modification, jeopardizing relationships or opportunities, and the DSM-IV-specific co-dependence (needing to borrow money to game) — and are listed in full in Table S2 (see also Table S6).

The Cronbach’s α for this scale was 0.70 (95% confidence interval obtained with bootstrapping [0.67 – 0.72]). The observed α, while in the lower acceptable range, is in line with values reported for the most widely used DSM-based gaming-disorder instruments in adolescent samples (e.g., α = 0.81–0.82 for the 9-item Internet Gaming Disorder Scale, [51]; α = 0.72 for the IGDT-10, [52]; α = 0.69–0.85 across 9 countries in the IGDT-10 cross-cultural validation, [53]; α = 0.56–0.78 across three German child/adolescent waves for the IGDS9-SF, [54]; see also [55]).

We further report McDonald’s ω for all multi-item scales throughout the manuscript as, unlike α, it does not assume tau-equivalence and is therefore more appropriate for brief DSM-derived symptom inventories like ours [56,57]. McDonald’s ω total for the same 10-item scale was 0.75, which is again in the low acceptable end of the range reported for adult IGDS9-SF / GDT validation samples and within the range observed in adolescent samples [54]. Finally, removing the single DSM-IV item not retained in the DSM-V IGD criteria (co-dependence) did not improve reliability (α = 0.69; ω = 0.74).

#### VG consumption

Participants who said they usually play video game were asked to report the average number of hours of gaming on a typical weekday and on a typical weekend day. An hour/week composite was derived by multiplying the number of weekday hours by five and adding the number of weekend hours multiplied by two.

#### Impulsiveness

The students’ level of impulsivity was measured using a shortened version of the Barratt Impulsiveness Scale [58]. Students rated each item on a four-point scale ranging from strongly disagree (1) to strongly agree (4). The scale included both positive worded items such as “I keep my feelings under control” and negative worded items such as “I talk even when I know I shouldn’t.” After reversing the positive worded items for consistency of direction of expression, a mean score of the total items was used to represent the level of impulse control problem, where a higher score indicates a higher level of impulse control problem. The Cronbach’s α for impulsiveness was 0.69 (95% CI = [0.66– 0.70]), and the McDonald’s ω was 0.76.

#### Family environment

This scale, which was adapted from Glezer’s (1984) instrument, measures perceptions of home living environment. Some items measure positive conditions (e.g. “It is pleasant living with my parents and family”) and others measure negative conditions (e.g. “Generally, there is nothing good about living at home”). Response is based on a 4-point rating scale, from “Strongly disagree” to “Strongly agree.” The responses to these questions were summed (after reverse-scoring when necessary), such that higher values correspond to more positive family environments. The Cronbach’s α for this the family environment scale was 0.78 [0.76 – 0.79] and the Mc Donal’s ω was 0.83.

The list of variables collected in the original study is available in Table 1 of Choo et al. 2010 [45] and the full list of papers published using this dataset is available in section 5 of the supplementary information. The proportion of missing data at baseline ranged from 0% for video game consumption (which was used for grouping) and 13% for impulsiveness; it also increased with each wave (see Table S1). The proportion of missing data was higher in occasional use gamers, except concerning gaming data at baseline (Figure S1). To reduce possible bias due to differential attrition between groups, our main analyses focused on completers, i.e., participants who had data at wave 1 and 3, and no missing-data imputation technique was applied. A comparison of completers and non-completers on baseline measures, as well as an analysis with multiple data imputation technique, indicated the same patterns as in the main analyses (see SI section 2 - Table S3, Table S4 and Figure S2).

**Table 1.** Participants*’* age, sex and education at Wave 1. * For continuous variables (Age), summary values correspond to mean (SD); For categorical variables (Sex, Education), summary values indicate the proportion (%) of the total sample.

| Variable* | N (%) | Age mean (SD) | Occasional use gamers | Low use gamers | Intermediate use gamers | Heavy use gamers |
| --- | --- | --- | --- | --- | --- | --- |
| **N** | 3034 | 11.20 (2.06) | 678 | 786 | 785 | 785 |
| **Sex** | 3408 |  |  |  |  |  |
| ... Male | 2500 (73%) | 11.20 (2.06) | 454 (69%) | 570 (73%) | 573 (73%) | 596 (76%) |
| ... Female | 908 (27%) | 11.20 (2.03) | 206 (31%) | 215 (27%) | 210 (27%) | 188 (24%) |
| **Education** | 3035 |  |  |  |  |  |
| ... 3rd grade | 743 (24%) | 8.75 (0.56) | 196 (29%) | 242 (31%) | 148 (19%) | 157 (20%) |
| ... 4th grade | 711 (23%) | 9.64 (0.53) | 140 (21%) | 253 (32%) | 186 (24%) | 132 (17%) |
| ... 7th grade | 916 (30%) | 12.60 (0.67) | 142 (21%) | 183 (23%) | 276 (35%) | 315 (40%) |
| ... 8th grade | 665 (22%) | 13.60 (0.66) | 200 (29%) | 108 (14%) | 175 (22%) | 181 (23%) |

Pearson correlation analysis (Figure S3) indicated that each change score was correlated positively with wave 3 data and negatively with wave 1 data. Video gaming time and impulsivity were positively correlated both within and across waves, whereas family environment correlated negatively with both impulsivity and video gaming time.

### Ethics

The study was carried out in accordance with relevant guidelines and regulations. Approval for this study was obtained from the ethics review board of the National Healthcare Group (Singapore) prior to the study commencement. Data collection took place over a three-month period from December to February of each school year between 2007 and 2009. All of the procedures and materials were approved by the Ministry of Education (MOE) and each of the participating schools. Formal permissions were granted by the respective school authorities to conduct surveys with their students, following local ethical guidelines. Once the study was approved by the MOE and each individual school, then the schools managed the consent process based on their policies, which could be different from school to school. Thus, for this study involving minors, informed consent was obtained from all participants or their legal guardians.

### Statistical analyses

To assess the impact of changes in VG consumption, impulsiveness and family environment on changes in PG, we first computed difference scores between wave 1 and 3 for PG, VG consumption, impulsiveness and family environment. We then regressed changes in PG with changes in VG consumption (weekly hours), and used Specification Curve Analysis (SCA) to adjust for all possible factors known to influence the outcome (PG) or its predictors (VG hours, impulsiveness, family environment) to ensure that the observed relationships cannot be explained by a third factor related to school, education level, race, sex, maternal education or paternal education.

We then conducted moderation analyses to test whether the relationship between gaming hours and impulsivity with PG were statistically moderated by gaming group. Finally, Cross-Lagged Panel models (CLPM) and Random-Intercept Cross-Lagged Panel Models (RICLPM) allowed us to test the direction of influences^1^. While the CLPM does not distinguish between inter-individual (stable between-person trait) and intra-individual (within-person changes) effects, the RI-CLPM addresses this limitation by decomposing the variance into a stable between-person random intercept and wave-specific within-person deviations, thereby isolating the purely reciprocal dynamics occurring over time. Unlike the SCA and moderation analyses, CLPM and RI-CLPM were run including T2 in addition to T1 and T3, but did not distinguish between gamers groups. Analyses were conducted in R (Version 4.6.1). Specification-curve analyses were implemented with the specr package; partial η² effect sizes and Type-III ANOVAs were computed with the car, effectsize and heplots packages; linear mixed-effects moderation models were fitted with lme4 (with afex, emmeans and interactions for follow-up tests); and the cross-lagged (CLPM) and random-intercept cross-lagged (RI-CLPM) panel models were estimated with lavaan using MLR estimator with FIML for missing data. Statistical significance was evaluated at α = .05, and effect sizes were interpreted using the conventional benchmarks reported above. Model fits were estimated using AIC and BIC for ANOVAs and mixed-effects models and with CFI, RMSEA and SRMR for RI-CLPM models.

## Results

The data from 3,434 adolescents were collected between 2007 and 2009 in Singaporean Primary 3 (corresponding to 3^rd^ grade, n = 743), Primary 4 (4^th^ grade, n = 711), Secondary 1 (7^th^ grade, n = 916) and Secondary 2 (8^th^ grade, n = 664) schools. The sample consisted of 2,500 males (73%) and 908 females (24%). The higher male percentage is due to three of the 12 participating schools being males only, and not differential participation rates. We did not use data from Wave 4 collected in 2010 (N = 973) because by that year about two-thirds of the children had changed schools (e.g., moving from elementary to secondary schools) resulting in a high drop-out rate which would have penalized the analysis of waves 1 - 3.

Each year, participants completed a number of surveys that included, among others, questionnaires assessing PG, video gaming habits, impulsiveness, and family environment. Information about racial makeup and SES of the whole sample can be found in previous publications [45]. The variables analysed in this study include self-reported measures of weekly hours of video game play (higher score indicates higher consumption), the Barratt Impulsiveness Scale (higher score = higher impuslivity), and the Glezer’s [59] family environment instrument (higher score = more supportive environment).

Sample description is available in Table 1 (wave 1). Occasional use gamers included a lower proportion of males (69%) compared to heavy use gamers (76%) with low use gamers (73%) and intermediate use gamers (73%) falling in-between. Both primary (3^rd^ and 4^th^ grades) and secondary (7^th^ and 8^th^ grades) students were well represented in each video game group with the proportion of children from primary, for example, being 50%, 63%, 43% and 37% in occasional use gamers, low use gamers, intermediate use gamers and heavy use gamers respectively.

Descriptive statistics for the primary outcome measure (PG) and the independent predictor variables (VG hours, impulsiveness and family environment) are summarized in Table 2. Average weekly gaming time of 20.5 hours per week was in the expected range for an adolescent population (ages 11-13) in these years (2007-2009). Note that as reported in Gentile et al. (2011) [47], PG decreased on average from wave 1 to wave 3. Here, we will be interested in modeling individual changes and not mean behavior. Missing data per wave and attrition rates per group are provided in supplementary Table S1 and Supplementary Table S2.

**Table 2.**
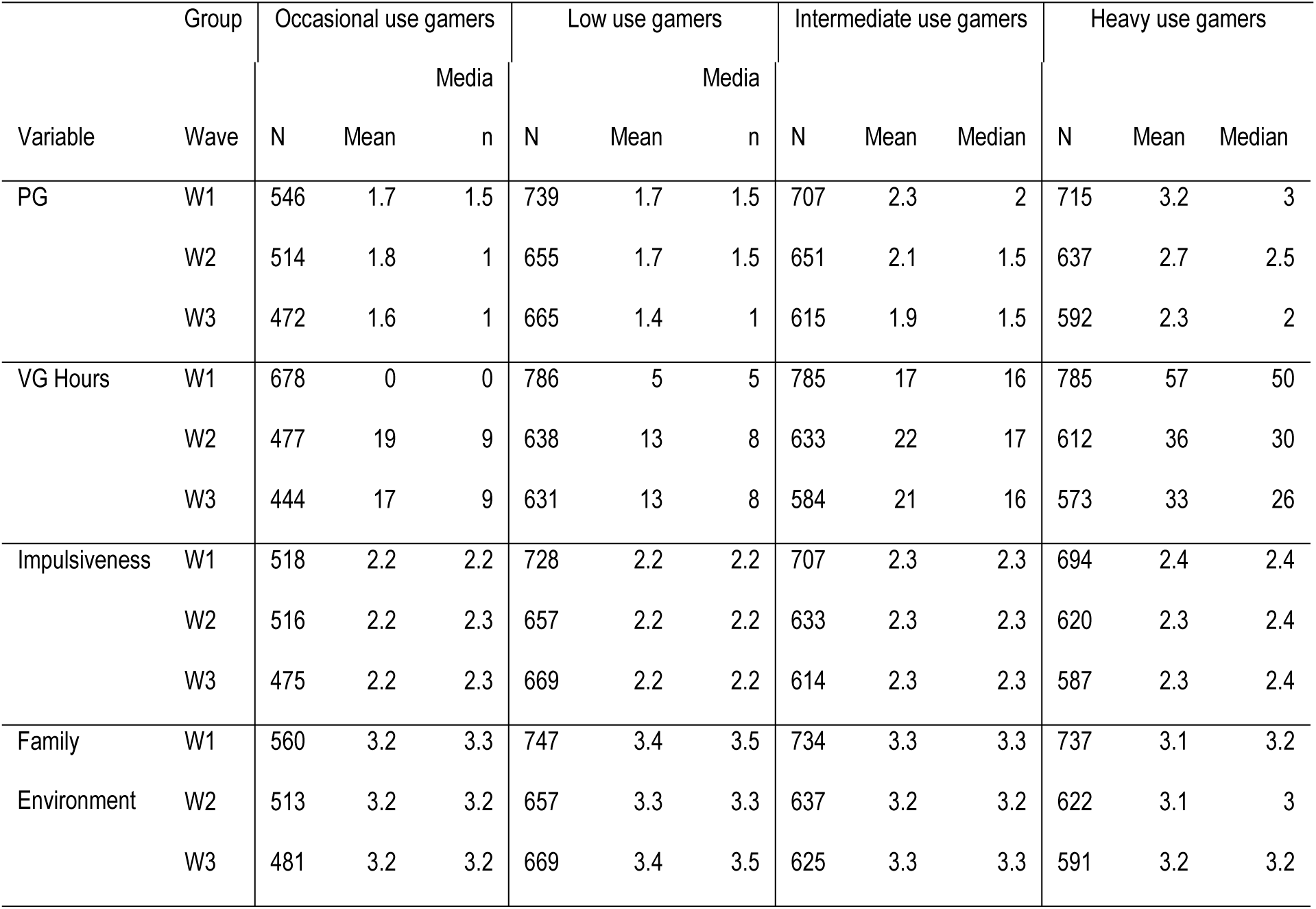
Descriptive data for the primary variables of interest across the 3 waves: Problematic Gaming (PG), weekly video gaming hours (VG Hours), impulsiveness, family environment. Wave = data collection period with W1 = 2007, W2 = 2008 and W3 = 2009.

To quantify the impact of *changes* in VG hours, impulsiveness and family environment on *changes* in PG, we ran separate regression analyses predicting changes in PG scores from change scores in VG hours, in impulsiveness, and in family environment during the same time period. Our primary analysis uses differences between wave 1 and wave 3. More granular analyses between waves 1 and 2 and then between waves 2 and 3 indicate similar trends, albeit in a more noisy fashion (see SI section 4.4, Figure S4).

To qualify different levels of video game consumption at study start, occasional use gamers, who reported not usually playing video games at study inception, were kept separate from other players who were further divided into 3 equal sized groups using quartiles of self-reported video game consumption at wave 1. In addition to occasional use gamers (N = 678), the three groups of players defined by this quantile analyses were low use gamers (N = 786) playing between 1 and 10 hours per week at study inception, intermediate use gamers (N = 785) playing 10-28 hours per week which is typical for adolescents of this age range, and heavy use amers (N = 785) playing on average 57 hours per week, but up to 106 hours for the most avid players. This latter group aligns well with the 30 hours/week cutoff proposed by the APA [1] to differentiate problematic gamers (more than 30 h/w) from heavy use gamers (less than 30h/w). Importantly, sample size in all four groups is well matched.

Figure 1 shows the distribution of all variables at wave 1 and wave 3, with each grey line showing within-participants’ changes. Note that occasional use gamers reported some PG symptoms at wave 1 despite reporting not *usually* playing video games. Although this may appear surprising, it likely reflects differences in the wording of our question. Four sets of counterbalanced questionnaires were used. All questions including those regarding gaming behavior and PG symptoms specified the time interval to be “over the past year.” Children were therefore asked if, during the past year, they usually played video games. Gamers that replied “No,” or occasional use gamers as we labeled them, exhibited levels of PG symptoms at wave 1 as high as at wave 3, and nearly as high as all other groups at baseline. Occasional use gamers - while signaling they don’t usually play at present - may have played regularly in the past, but stopped due to encountering difficulties, or may be children who were not allowed to play video game at home, and yet may engage in video game play outside of the family setting, signaling an attitude around gaming, which is again rather unhealthy. In either case, these occasional use gamers may report on experienced past symptoms. This apparent disconnect aligns with their most frequently reported “symptoms” at baseline being loss of control / unsuccessful attempts (item 9), followed by escape (item 6).

**Figure 1.**
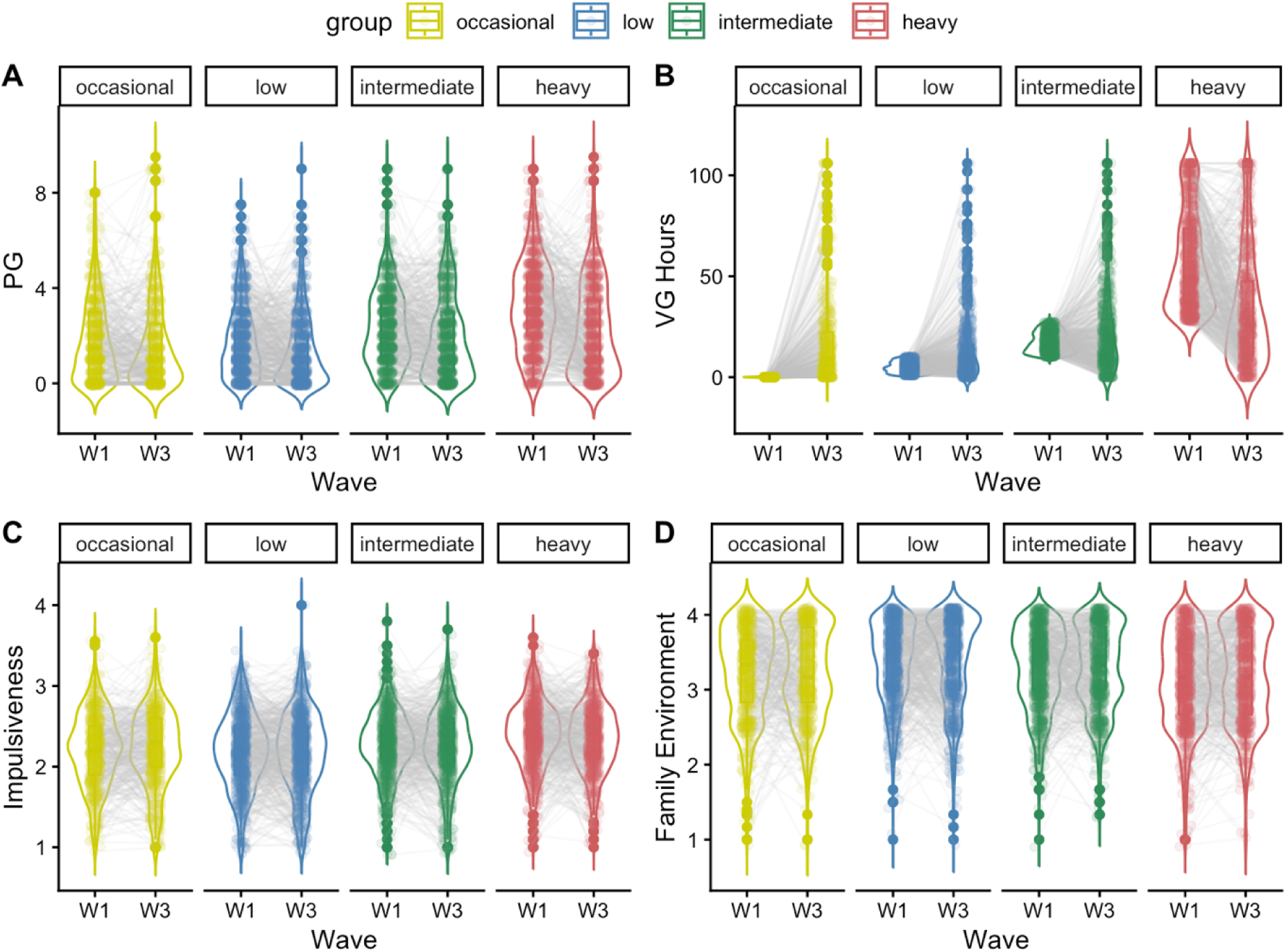
Distributions of scores at wave 1 (W1) and wave 3 (W3) for PG (A), VG hours (B), impulsiveness (C) and family environment (D) in occasional use gamers, low use gamers, intermediate use gamers and heavy use gamers. Sample sizes at wave 1 were 2434 for PG, 3000 for VG hours, 2933 for impulsiveness and 2931 for family environment; Sample sizes at wave 3 were 1872 for PG, 2634 for VG hours, 2438 for impulsiveness and 2343 for family environment. From wave 1 to wave 3, PG decreased in all groups, VG hours increased in occasional use gamers, low use gamers and intermediate use gamers and decreased in heavy use gamers. Impulsiveness and family environment did not change between wave 1 and wave 3. Descriptive statistics of the scores across waves and groups are available in SI Section 3.1.

To isolate the effect of change in predictors on the change of outcome, each analysis controlled for the absolute value of both outcome (i.e., PG) and the predictor (e.g., VG hours, impulsiveness, and family environment) at study start. In doing so, we ensure that the estimated effects are really due to changes in predictors and not to the absolute level of our measurements at the beginning of the study. Controlling VG hours at study inception —which defines our gaming groups— ensures that changes in VG hours and not any baseline difference carries the effect of interest. Importantly, the impact of a change in VG hours on changes in PG was the same whether controlling for VG hours at study inception (Figure 2) or not (Figure S3). In addition, we used specification curve analysis (SCA) to adjust for all possible combinations of the most common confounding factors, namely age, sex, race, education, parental education (father and mother) and school. Using SCA allows to draw robust inferences on the association between two variables while controlling for other possible sources of confounding influence [60]. In this analysis, any effect size (partial eta squared) at or below .01 is considered negligible, those between .01 and.06 as small, between .06 and .14 as medium and above .14 as large [61].

**Figure 2.**
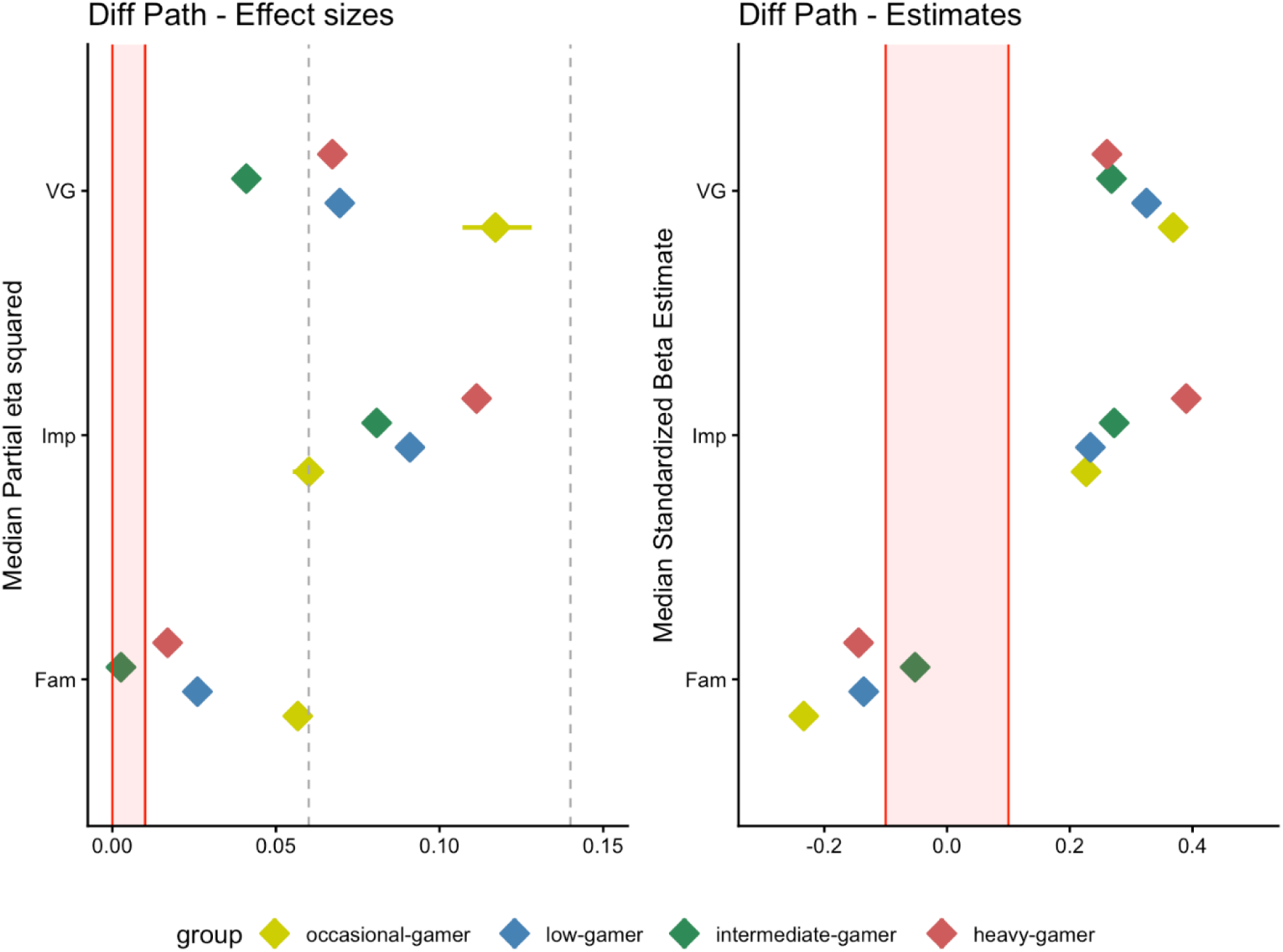
Results of the SCA analysis based on change scores between wave 1 and wave 3. Left panel: partial eta-squared which reflect the % of explained variance in PG changes by changes in each predictor. Right panel: standardized beta coefficients showing the direction and strength of the association. VG = changes in weekly VG hours, Imp = changes in impulsiveness, Fam = changes in family environment.

Effect sizes and regression coefficients are available in Figure 2 and Table S2. In occasional use gamers, a change in PG between wave 1 and 3 was best explained by a change in VG hours, controlling for PG score and VG hours at study inception. More specifically, in occasional use gamers, 12% of the interindividual variability in PG change between waves 1 and 3 is explained by interindividual change in VG hours between waves 1 and 3. Strikingly in heavy use gamers, 11% of the variability in PG change between waves 1 and 3 is explained instead by change in impulsivity between waves 1 and 3, with a change in VG hours explaining less variability in PG change (7%). Overall, everything remaining equal, an increase in VG hours had a small-to-medium impact on PG change in all gaming groups, except for occasional use gamers where the effect size was clearly medium. In contrast, increases in impulsiveness had a medium impact on PG increase in all gaming groups, except in the occasional-gamers where the effect of increases in impulsiveness on PG increase was only small-to-medium. Finally, improvements in family environment were associated with decreases in PG, but only with small-to-medium effect sizes at best (at most, 6% of the variance in PG decrease is explained by family environment improvement in occasional use gamers). Overall, our results suggest that while changes in VG hours affect changes in PG in occasional use gamers more strongly than in heavy use gamers, changes in impulse control instead affect changes in PG in heavy use gamers more strongly than occasional use gamers. This pattern suggests that baseline gaming habits (i.e., VG hours at wave 1) are critical in better understanding the impact of risk and predictive factors on the evolution of PG.

To further test whether changes in our two risk factors – VG hours and impulsivity – indeed interacted with baseline gaming status (i.e., VG hours at inception) in predicting PG change, we ran a Linear Mixed Model testing specifically the interaction between our two extreme gamer groups (occasional use gamers/heavy use gamers) and changes in VG hours (respectively changes in impulsiveness) in predicting change in PG. Each analysis was conducted while controlling for levels of PG, VG hours, and Impulsiveness at wave 1, as well as all other control variables (i.e., school, education level, race, sex, maternal education or paternal education).

As expected, the values for PG at wave 1 accounted for most of the variance in changes in PG from wave 1 to 3 (39%). In contrast, VG hours, impulsiveness, and sex at wave 1 had only a negligible-to-small effect, each accounting for about 1% of the PG change variability (Table 4). Most importantly, the two interactions, between gamers group and changes in VG hours as well as between gamers group and changes in impulsiveness, accounted each for 8% of the variance in PG change (Table 3). Note that this is a very conservative estimate, because by controlling for initial PG symptoms, we are also controlling for everything that explains or causes that initial level too.

**Table 3.** Coefficients and effect sizes for the effect of a change in video game consumption, impulsiveness and family environment, on changes in PG, from wave 1 to wave 3 (all reported after controlling for school, education level, race, sex, maternal education or paternal education as well as PG at wave 1).

| Predictor | Group | Partial eta squared | Beta | Std.Dev | P value | Observations |
| --- | --- | --- | --- | --- | --- | --- |
| VG Hours | Occasional Use Gamers | 0.117 | 0.369 | 0.063 | <0.001 | 275 |
|  | Low Use Gamers | 0.069 | 0.325 | 0.051 | <0.001 | 567 |
|  | Intermediate Use Gamers | 0.041 | 0.268 | 0.059 | <0.001 | 495.5 |
|  | Heavy Use Gamers | 0.067 | 0.261 | 0.046 | <0.001 | 479.5 |
| Impulsiveness | Occasional Use Gamers | 0.060 | 0.227 | 0.055 | <0.001 | 283 |
|  | Low Use Gamers | 0.091 | 0.234 | 0.031 | <0.001 | 584.5 |
|  | Intermediate Use Gamers | 0.081 | 0.272 | 0.042 | <0.001 | 503 |
|  | Heavy Use Gamers | 0.111 | 0.390 | 0.052 | <0.001 | 464.5 |
| Family Environment | Occasional Use Gamers | 0.057 | -0.234 | 0.061 | <0.001 | 269.5 |
|  | Low Use Gamers | 0.026 | -0.136 | 0.035 | <0.001 | 594.5 |
|  | Intermediate Use Gamers | 0.003 | -0.052 | 0.045 | 0.26 | 516.5 |
|  | Heavy Use Gamers | 0.017 | -0.144 | 0.051 | 0.005 | 483.5 |

**Table 4.** Results of the linear model with Δ*PG* as depedent variable.

|  | *Sum* |  |  |  | *Eta squared* |
| --- | --- | --- | --- | --- | --- |
|  | *Sq* | *Df* | *F value* | *Pr(>F)* |  |
| **W1_PG** | **264.80** | **1** | **459.07** | **< .001** | **0.39** |
| **W1_VG** | **26.72** | **1** | **46.33** | **< .001** | **0.01** |
| **W1_Impulsiveness** | **17.66** | **1** | **30.62** | **< .001** | **0.00** |
| Age | 0.12 | 1 | 0.21 | 0.65 | 0.00 |
| Race | 0.06 | 1 | 0.11 | 0.74 | 0.00 |
| **Sex** | **7.17** | **1** | **12.42** | **< .001** | **0.01** |
| School | 0.62 | 1 | 1.07 | 0.30 | 0.00 |
| Education | 0.13 | 1 | 0.22 | 0.64 | 0.00 |
| Maternal education | 0.16 | 1 | 0.28 | 0.60 | 0.00 |
| Paternal education | 0.28 | 1 | 0.48 | 0.49 | 0.00 |
| **$\Delta VG \times Group$** | **28.80** | **2** | **24.97** | **< .001** | **0.08** |
| **$\Delta Impulsiveness \times$** | **35.20** | **2** | **30.51** | **< .001** |  |
| **Group** |  |  |  |  | **0.08** |
| Residuals | 430.10 | 638 |  |  |  |

Although the APA diagnosis of PG rests on observing more than 5 symptoms out of 9, it is possible to reach that threshold through rather different combinations of symptoms. To further explore how changes in gaming time may relate to change in individual symptoms of the PG scale, especially in occasional use gamers, we ran an exploratory analysis assessing how changes in gaming hours predict changes in individual items of the PG scale (SI section 5). Figure 3 shows the effect sizes of the associations (again controlled for VG hours and PG score at study inception). Changes in VG hours had an impact on changes of the sneaky (8.4%) and deception (6.5%) symptoms in occasional use gamers by a medium effect size, while only having negligible effects in the three other gaming groups (< 1.3%). The other changes in symptoms that were moderately associated with changes in VG hours in occasional use gamers, as well as in other gaming groups, were tolerance (12.1%) and preoccupation (7.1%). The remaining effects, seen on change in escape, loss of interest, loss of control and withdrawal were small in size for all groups (< 5.2%). Although in need of replication as these analyses are exploratory, the possibility that the occasional-gamers may be more likely to increase in jeopardizing and deception symptoms as their VG hours increase is in-line with the known psychological burden of hiding one’s playing if it might be seen as interfering with other valued goals.

**Figure 3.**
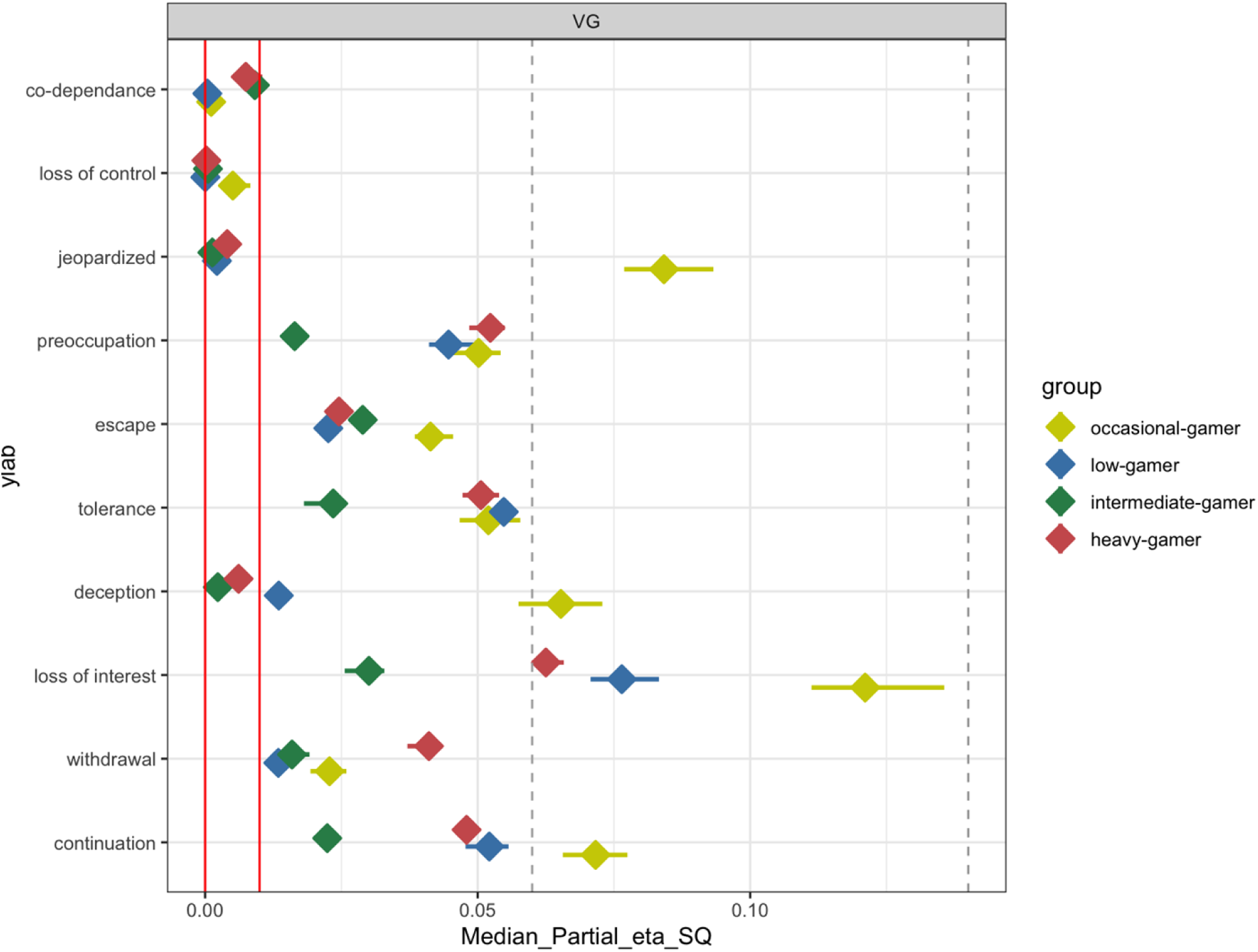
Effect sizes (partial eta-squared) of the impact of changes in gaming behavior (VG hours) on changes in each PG symptom over 2 years (W1-W3).

Analyses of difference scores do not inform about the moderation effects or about the directionality of influences. Therefore, additional exploratory analyses presented in the supplementary material were conducted to more formally test the moderating influence of VG hours and impulsivity on PG as well as the directionality of the associations of VG consumption, Impulsivity, and family environment with PG. Consistent with our main results, the moderating influence of VG hours and impulsivity were also confirmed. Interactions were found between gaming time and group (F(3,117.5) = 10.9, p < .001) and impulsivity and group (F(3,41.3) = 4.0, p < .01). The impact of VG hours on PG was stronger in occasional us egamers, whereas the effect of impulsivity was stronger in heavy use gamers (Table S3 and Figure S5).

Cross-lagged panel results (Table S3 and Figure S6) provided complementary results, showing significant longitudinal associations at the within-person level only, and none at the between-person level. Concerning within-person level, no significant association was found between video gaming hours and PG. In contrast, a unidirectional (positive) influence of PG on impulsiveness was observed, where an increase in PG above baseline predicted an increase in impulsiveness the next year. Finally, these analyses also revealed bidirectional (negative) associations between family environment and PG, opening the door to multiple complex spirals of influence where (a) a degradation of family environment could predict later increase in PG symptoms, and (b) an increase in PG would further impair family environment (and increase impulsiveness). These within-person effects indicate that changes in one predictor relative to baseline were significantly related to changes in PG at the next time point. Yet, the absence of between-person influences suggests that individuals who scored high on a given trait-level predictor (VG hours, impulsiveness or family environment) were not higher (or lower) on PG across the study period.

## Discussion

Understanding the determinants of PG is of public health importance. Previous work has identified several risk and protective factors that are potentially modifiable through interventions. Yet, there remains large heterogeneity. Here, we investigated whether considering individual levels of video game consumption at study inception could help better characterize how naturally occurring longitudinal changes in such factors may be related to changes in PG.

The first noteworthy finding is a strong impact of intrapersonal factors. Changes in VG hours and in impulsivity explained between 4% and 12% of the changes in PG symptoms. Concerning interpersonal factors, we only had access to changes in family environment and these alone accounted for up to 6% of the variance in PG changes. As expected, the effects were in opposite directions with family environment acting as protective against PG, whereas VG hours and impulsivity acted as risk factors. These results echo those observed in meta-analytic works that systematically point to negative impacts of gaming consumption (VG hours) and impulse regulation issues (impulsiveness) on PG as compared to possible protective interpersonel or family environment ones [13].

More strikingly, the effects of video game hours and impulsivity on PG varied as a function of video gaming habits at study start. Changes in video game hours between waves 1 and 3 explained 12% of the variance in PG in occasional gamers (respectively 7% in heavy gamers). Conversely, changes in impulsivity explained 11% of the variance in PG in heavy use gamers (respectively 6% in occasional use gamers). Clearly, initial gaming status majorly impacts which intra-personal factors link to PG, calling for different approaches across gaming groups when it comes to better help them cope with possible PG symptoms.

In heavy use gamers, weak impulse regulation is a significant PG risk factor, numerically stronger than video game hours. Heavy use gamers may struggle to resist the urge to play when exposed to gaming cues. As a result, heavy use gamers are likely to score higher on several items of the PG scale, including gaming to escape, withdrawal and preoccupation, as well as loss of control (unsuccessful attempts to reduce gaming time). This would also align with PG symptoms being correlated with greater ADHD symptomatology [10,62]. ADHD symptoms early on at 6 years of age were found to predict PG at age 7, but then PG at 8 years of age was observed to predict ADHD symptoms at age 10. This is in-line with other studies documenting bi-directional links between ADHD and PG [63,64], and further suggests a possibly positive feedback loop [65]. Indeed, poor inhibitory control could exacerbate PG early in development, which in turn may lead to greater ADHD symptomatology later in development.

Whether PG may causally induce weaker impulse regulation remains, however, a hotly debated topic. In the field of violent VG play, reduced inhibitory control has been documented following playing a violent VG, lasting for tens of minutes after the end of play [66,67]. In line with such a causal view, neuroimaging work also indicates that a short (violent vs non-violent) gaming bout alters subsequent prefrontal activation as measured by inhibitory tasks [27,68]. Yet, whether such short-term acute exposure effects may result in long-term effects, such as the weaker impulse regulation seen here across two years, remains debated [69–72]. It is also unclear whether reduced inhibitory control may be witnessed after playing any game genre, or rather remains specific to just playing violent video games. While certainly an important topic of research for the future, by highlighting the unique association between poor inhibitory control and PG in heavy use gamers, the present work points to gamers’ characteristics as a key variable to better buffer the heterogeneity in studies of the link between PG and impulsivity.

Another point concerns the nature of the inhibitory control construct under study when looking at PG and impulsivity. A recent meta-analysis of 43 cross-sectional studies of disordered screen use [73] reported small-to-medium cognitive deficits relative to controls (overall Hedges’ g = 0.38), with attention being the most affected domain (g = 0.50) and the broad executive-functioning domain showing a smaller but significant deficit (g = 0.31). Within executive functioning, the strongest signals came from delay discounting (g = 0.59) and Stop-Signal-task performance (g = 0.52), while Go/No-go reaction times did not differ between disordered users and controls — indicating that the relationship between PG and inhibitory control is task-dependent rather than uniform. A complementary meta-analysis specifically targeting response inhibition in IGD reached a similar conclusion [74], and an independent meta-analysis of cognitive deficits in problematic internet use likewise reports the strongest deficits in inhibitory control and decision-making [75]. Two caveats are important. First, the available evidence is overwhelmingly cross-sectional, so it cannot, on its own, establish whether PG induces these deficits or whether pre-existing deficits predispose to PG — a question for which longitudinal designs such as ours are well-suited. Second, the trait-level impulsivity assessed by the BIS (used here) and the task-level inhibitory control captured in most meta-analyses are conceptually and psychometrically distinct [23]. Taken together, these results suggest that PG may not necessarily change trait impulsivity itself but may be accompanied by — and possibly amplify — difficulties in task-level inhibitory control, which our self-report measure of impulse-control problems would tend to pick up as elevated impulsiveness over time. This is consistent with our finding of bi-directional change-on-change effects between impulsivity and PG specifically in heavy use gamers.

Finally, our heavy use gamer category, defined solely on the basis of self-reported weekly hours (mean ≈ 57 h/week, up to 106 h/week), is likely composed of a mix of profiles, ranging from adolescents engaging in highly committed, skill-oriented, or competitive play to adolescents whose extensive use reflects difficulties in regulation of, and emerging problems with gaming. Accumulating evidence indicates that these profiles are not interchangeable. In a systematic review of brain-imaging studies, Choi et al., (2021) [76] showed that, despite comparable extensive engagement, professional/elite gamers and individuals with Gaming Disorder display distinct structural and functional alterations. Pro-gamers exhibit enhanced attentional and sensorimotor function as well as better cognitive control, whereas individuals with Gaming Disorder show impaired cognitive control and increased craving-related activation. A complementary line of work led by Billieux and colleagues has long argued for separating high engagement from problematic involvement and has shown that time spent playing, on its own, is a poor proxy for harm [77,78]; see also [79]. The fact that, in our data, changes in VG hours explained a smaller share of variance in PG among heavy use gamers (∼7%) than among occasional use gamers (∼12%) is consistent with this view. That is, if a non-negligible subset of heavy use gamers are highly engaged but not symptomatic, additional hours in this subgroup will be only weakly coupled to PG, attenuating the average VG-hours-to-PG association. Future work using measures that capture the motivational, functional, and contextual aspects of play (e.g., passion, achievement, social, competitive motives; functional impairment; subjective control) in addition to gaming time will be needed to disentangle these distinct trajectories within heavy use gamers.

When it comes to occasional use gamers, the present work indicates that resources may be better spent on addressing their growing interests in gaming activities. The impact of video game consumption on PG in occasional use amers was seen primarily in increases in jeopardizing behavior (stealing video game or money) and deception, as well as loss-of-interest and continuation. As development proceeds, beginning gaming activities may more regularly lead to continuation and in turn to loss-of-interest in other activities. Jeopardizing behaviors and deception may furthermore reflect attempts to better integrate this new behavior in one’s life without causing or signaling changes in social and family relationships. Such emerging functional difficulties during development may be particularly notable among more vulnerable individuals, such as those with low baseline impulse control or unsupportive family environments, feeding a positive feedback loop as described in [65]. Although analyses performed at the symptoms level of PG are exploratory, they highlight how future work may better address the developmetal time-course of symptoms emergence and their associations with changes in intra- or interpersonal factors by modeling how gaming habits evolve from study start.

By leveraging a large, longitudinal dataset combined with a novel stratification approach, this study addresses the heterogeneity of associations between problematic gaming and intra- and inter-personal factors reported in the literature. Our analytical approach combined the strengths of multiple approaches. First, SCA results suggest associations between PG and VG hours in occasional use gamers, between PG and impulsivity in heavy use gamers as well as between PG and family environment even when controlling for possible confounding impact of age, gender, school and parental education. Second, moderation analyses confirmed that the gaming group (i.e., baseline gaming consumption) moderated the longitudinal associations of gaming time and of impulsiveness with PG. Third, CLPM and RICLPM allowed us to disentangle the direction of influences, revealing (a) no significant influences when considering mean trends (i.e., adolescents scoring always high or low on a given trait didn’t always score high or low on PG), whereas significant associations were found at the intra-individual level (i.e., considering within-person temporary state increases or decreases). More specifically, we found (b) a unidirectional within-person (positive) impact of PG on impulsiveness, and (c) a bidirectional (negative) relation between PG and family environment. Finally, the small-to-moderate magnitude of the observed effects compares well with previous research [9,19,20]. Our beta coefficients of 0.37 (impact of VG hours on PG in occasional use gamers) and 0.39 (bi-directional associations between impulsivity and PG in heavy use gamers), and the fact that these explained variances 11-12% of the variance in PG suggest practical significance. Indeed, an R² of 0.12 is considered a small-to-moderate effect in behavioral sciences. It is comparable in strength to the associations between screen time and mental health in longitudinal studies which ranges from 0.05 to 0.15 for single predictors [80]. For comparison, effects of physical activity predicting BMI over similar time spans are also comparable in magnitude (R² ≈ 0.10–0.20, [81]).

A few methodological limitations are worth mentioning. First, results of cross-lagged (CLPM and RI-CLPM) analyses, as well as symptom-specific associations, should be considered exploratory and in need of replication. Second, among our measures, family environment aimed to capture different aspects of parental behaviour and parent–child relationships. The original cohort was not designed to assess peer- or school-level dynamics (validated measures of school belonging, peer victimization or peer influence were not collected). A substantial body of work — including studies in Hong Kong [82], mainland China [83,84] and broader East Asian adolescent samples (for syntheses see [11,34,85]) — has documented that peer victimization, peer influence and school engagement are robust risk and protective factors for problematic gaming, sometimes accounting for variance beyond that explained by the family environment. Family environment in our analyses is therefore best interpreted as one facet of the broader interpersonal layer, and our estimate of its contribution to interpersonal factors should be regarded as a lower bound. Future longitudinal work in adolescents would benefit of incorporating peer- and school-level measures alongside family functioning to allow direct comparison of their relative contributions to changes in PG over time.

Third, our gaming-group categorization rests on self-reported weekly hours and does not distinguish between healthy high engagement (e.g., competitive, passionate, or skill-oriented play) and problematic involvement. As a consequence, the heavy use gamer group is intrinsically heterogeneous and likely includes adolescents whose extensive play does not reflect disorder. This is expected to attenuate, rather than inflate, the observed effect of VG hours on PG in that group, and underscores the value of complementing hours-based measures with indicators of motivation, functional impairment, and subjective control in future work. We emphasize that these labels (occasional, low, intermediate, heavy) denote positions along a continuum of self-reported engagement rather than discrete or clinically meaningful categories.

Fourth, the impulsiveness questions we used, albeit coming from a commonly used instrument, measured impulse control problems, reflecting the behavioral impact of impulsivity. As such, it may not capture the multiple dimensions of the impulsivity construct per se. Future work should distinguish between the psychological trait impulsivity, which corresponds to the multidimensional tendency to act prematurely, without foresight or adequate deliberation, and state impulsiveness, which corresponds to the observable behavioral consequences of impulsive actions in the moment.

Fifth, the present measure of gaming consumption is based on typical-day composite (typical weekday × 5 + typical weekend day × 2), which can overestimate weekly hours for participants who do not play every day and is, more broadly, an imperfect index of harm. Across screening and assessment instruments, gaming time correlates only modestly with gaming-disorder symptom scores (largely r ≈ 0.2–0.4; see the systematic review by King et al., 2020 [86]), and a recent psychometric evaluation indicates that self-reported gaming-time measures track available leisure time more closely than they track problematic patterns of play [87]. This is consistent with our finding that changes in VG hours account for a relatively modest share of variance in PG change. This point is particularly relevant for the interpretation of the larger VG-hours → PG association observed in occasional use gamers, as partly reflecting baseline differences in leisure-time availability and engagement rather than a specific symptomatic intensification. As many others have pointed out, more objective measures of game consumption would be preferable. In particular, structured retrospective methods such as the Timeline Followback (TLFB), behavioural-frequency approaches, and objective indices such as platform game-logs (telemetry) and data-donation designs have been used to obtain more accurate estimates of gaming time and could mitigate these concerns in future work (e.g., [88]). Finally, we recognize the gaming landscape has changed substantially since our data set was collected, particularly in the prevalence of mobile gaming, online multiplayer formats, and free-to-play monetization models. The findings from this historical cohort should thus be extrapolated to contemporary gaming with caution.

In sum, the present work informs interventions designed at attenuating risk-factors and promoting protective factors, by advocating for a differential approach as a function of gaming consumption. Our results suggest that interventions designed to boost impulse control in adolescents may be most helpful to reduce PG symptoms in heavy use gamers. Conversely, when it comes to occasional use gamers, interventions may benefit from being designed around the disconnect between their growing motivation to play and the social consequences of this new activity for them. Thus, while time reduction strategies are often first implemented, multi-component programs addressing all layers from the agent (game consumption), the host (impulsivity) and the environment (family environment), will likely be more effective. In that respect, cognitive behavioral therapy (CBT) with its more comprehensive approach, appears as a promising tool. In adults, the impact of CBT on gaming disorder is of moderate to large magnitude with effect sizes (Hedges’ g) between g = 0.72 [89] and g = 1.00 [90]. In adolescents, a moderate impact of CBT on gaming disorder symptoms has been reported of Hedge’s g = 0.55, or about half a standard deviation [91]. Such results point to the importance of education programs around healthy video game use (and for that matter digital media consumption), which remains unfortunately scarce [92], rather than on policy interventions focusing solely on playtime limits.

### Conclusion

By stratifying adolescents according to their baseline gaming habits, this study shows that the factors most strongly linked to changes in problematic gaming differ across engagement levels: changes in gaming time dominate in occasional use gamers, whereas changes in impulse control dominate in heavy use gamers. These findings argue against a one-size-fits-all approach and support tailoring prevention and intervention to a young person’s current gaming profile —suggesting that impulse regulation in heavy use gamers, and addressing the social integration of an emerging interest in gaming among occasional use gamers could be meaningful interventions. More broadly, they underscore that time spent playing is, on its own, an incomplete index of harm, and that future work combining engagement level with measures of motivation and function will be needed to refine these recommendations.

## Data availability

The data cannot be made publicly available due to ethical considerations involving Singaporean children and agreements with the funding agencies. The created summary variables are available upon reasonable request to the Research Support Office at Nanyang Technological University, NTU Singapore. The funders had no role in study design, data collection and analysis, decision to publish, or preparation of the manuscript.

## Supporting Information

Supplementary methods contains additional details about the methods as well as the following tables and figures:

**Table S1. Proportion of missing data per measure and wave.**

**Table S2. Number of observations available for the different variables and waves (and difference between wave 1 and wave 3) in the raw dataset.**

**Table S3. Completers vs non-completers - continuous variables.**

**Table S4. Completers vs non-completers - categorical variables.**

**Table S5. Summary results of CLPM and RI-CLPM models using Problematic Gaming (PG) as the outcome (y) and video game consumption (VG), impulsiveness (Imp) and Family environment (Fam) as predictors (x).**

**Table S6. Items of the DSM-IV and PG scale as compared to description of DSM-V for IGD diagnosis. The questions used in our study cover all 9 of symptoms of the DSM-V criteria for IGD. The only divergence concerns a tenth item on co-dependence based on the DSM-IV / PG which is absent from DSM-V / IGD.**

**Figure S1 Proportion of missing data per variable, group and wave. PG = problematic gaming, VG = VG hours, Imp = impulsiveness, Fam = Family environment.**

**Figure S2 Left: correlation matrix between all variables at Wave 1 and Wave 3. Right: correlation between changes in VG consumption, changes in impulsivity and changes in family environment between wave 1 and wave 3.**

**Figure S3 Results of the analysis based on change scores between wave 1 and wave 3 (W1-W3) without controlling for VG at W1. Left panel: partial eta-squared which reflect the % of explained variance in PG changes by changes in each predictor. Right panel: standardized beta coefficients showing the direction and strength of the association**. Facets represent different predictors: VG = changes in weekly VG hours.

**Figure S4 Results of the analysis based on change scores between wave 1 and wave 2 (W1-W2) and between wave 2 and wave 3 (W2-W3). Left panel: partial eta-squared which reflect the % of explained variance in PG changes by changes in each predictor. Right panel: standardized beta coefficients showing the direction and strength of the association**. Facets represent different predictors: VG = changes in weekly VG hours, Imp = changes in impulsiveness, Fam = changes in family environment.

**Figure S5. Results of the moderation analysis showing an interaction effects between PG and gaming time (top) and between PG and impulsivity (bottom).**

## Footnotes

1 We thank our reviewers for suggesting these further analyses.

## References

1. American Psychiatric Association. Diagnostic and statistical manual of mental disorders: DSM-5. Washington, D. C.: American Psychiatric Association.; 2013.

2. Torres-Rodríguez A, Griffiths MD, Carbonell X, Oberst U. Internet gaming disorder in adolescence: Psychological characteristics of a clinical sample. J Behav Addict. 2018 Sep 1;7(3):707–18. doi:10.1556/2006.7.2018.75 PubMed PMID: 30264606; PubMed Central PMCID: PMC6426364.

3. World Health Orgnanization. International Classification of Diseases for mortality and morbidity statistics (11th Revision) [Internet]. 2018 [cited 2025 Nov 21]. Available from: https://www.who.int/standards/classifications/classification-of-diseases

4. Reed GM, First MB, Kogan CS, Hyman SE, Gureje O, Gaebel W, et al. Innovations and changes in the ICD-11 classification of mental, behavioural and neurodevelopmental disorders. World Psychiatry. 2019 Feb;18(1):3–19. doi:10.1002/wps.20611 PubMed PMID: 30600616; PubMed Central PMCID: PMC6313247.

5. Darvesh N, Radhakrishnan A, Lachance CC, Nincic V, Sharpe JP, Ghassemi M, et al. Exploring the prevalence of gaming disorder and Internet gaming disorder: a rapid scoping review. Systematic Reviews. 2020 Apr 2;9(1):1. doi:10.1186/s13643-020-01329-2

6. Rehbein F, Gentile DA, Lemmens J, Rumpf HJ. Structural Game Characteristics and Problematic Gaming. SUCHT [Internet]. 2024 Apr 9 [cited 2025 Dec 2]. Located at: world. Available from: https://econtent.hogrefe.com/doi/10.1024/0939-5911/a000859

7. King DL, Delfabbro PH, Perales JC, Deleuze J, Király O, Krossbakken E, et al. Maladaptive player-game relationships in problematic gaming and gaming disorder: A systematic review. Clinical Psychology Review. 2019 Nov 1;73:101777. doi:10.1016/j.cpr.2019.101777

8. King D, Koster E, Billieux J. Study what makes games addictive. Nature. 2019;573(7774):346. doi:10.1038/d41586-019-02776-1

9. Gao YX, Wang JY, Dong GH. The prevalence and possible risk factors of internet gaming disorder among adolescents and young adults: Systematic reviews and meta-analyses. Journal of Psychiatric Research. 2022 Oct 1;154:35–43. doi:10.1016/j.jpsychires.2022.06.049

10. Koncz P, Demetrovics Z, Takacs ZK, Griffiths MD, Nagy T, Király O. The emerging evidence on the association between symptoms of ADHD and gaming disorder: A systematic review and meta-analysis. Clinical Psychology Review. 2023 Dec 1;106:102343. doi:10.1016/j.cpr.2023.102343

11. Schneider LA, King DL, Delfabbro PH. Family factors in adolescent problematic Internet gaming: A systematic review. Journal of Behavioral Addictions. 2017 Sep 1;6(3):321–33. doi:10.1556/2006.6.2017.035

12. Coşa IM, Dobrean A, Georgescu RD, Păsărelu CR. Parental behaviors associated with internet gaming disorder in children and adolescents: A quantitative meta-analysis. Curr Psychol. 2023 Aug 1;42(22):19401–18. doi:10.1007/s12144-022-04018-6

13. Zhuang X, Zhang Y, Tang X, Ng TK, Lin J, Yang X. Longitudinal modifiable risk and protective factors of internet gaming disorder: A systematic review and meta-analysis. J Behav Addict. 2023 Jun 29;12(2):375–92. doi:10.1556/2006.2023.00017 PubMed PMID: 37224007; PubMed Central PMCID: PMC10316169.

14. Li L, Niu Z, Song Y, Griffiths MD, Wen H, Yu Z, et al. Relationships Between Gaming Disorder, Risk Factors, and Protective Factors Among a Sample of Chinese University Students Utilizing a Network Perspective. Int J Ment Health Addiction. 2024 Oct 1;22(5):3283–301. doi:10.1007/s11469-023-01049-3

15. Jeong H, Yim HW, Lee SY, Lee HK, Potenza MN, Lee H. Factors associated with severity, incidence or persistence of internet gaming disorder in children and adolescents: a 2-year longitudinal study. Addiction. 2021;116(7):1828–38. doi:10.1111/add.15366

16. Orth U, Meier LL, Bühler JL, Dapp LC, Krauss S, Messerli D, et al. Effect size guidelines for cross-lagged effects. Psychological Methods. 2024;29(2):421–33. doi:10.1037/met0000499

17. Eichenbaum A, Kattner F, Bradford D, Gentile DA, Green CS. Role-playing and real-time strategy games associated with greater probability of Internet gaming disorder. Cyberpsychol Behav Soc Netw. 2015;18(8):480–5. doi:10.1089/cyber.2015.0092

18. Eichenbaum A, Kattner F, Bradford D, Gentile DA, Choo H, Chen V, et al. The role of game genres and the development of internet gaming disorder in school-aged children. Journal of Addictive Behaviors,Therapy & Rehabilitation. 2015 Sep 1;4(3). doi:10.4172/2324-9005.1000141

19. Na E, Choi I, Lee TH, Lee H, Rho MJ, Cho H, et al. The influence of game genre on Internet gaming disorder. J Behav Addict. 6(2):248–55. doi:10.1556/2006.6.2017.033 PubMed PMID: 28658960; PubMed Central PMCID: PMC5520129.

20. Liao Z, Chen X, Huang S, Huang Q, Lin S, Li Y, et al. Exploring the associated characteristics of Internet gaming disorder from the perspective of various game genres. Front Psychiatry. 2023 Jan 12;13:1103816. doi:10.3389/fpsyt.2022.1103816 PubMed PMID: 36713922; PubMed Central PMCID: PMC9878381.

21. Zha R, Tao R, Kong Q, Li H, Liu Y, Huang R, et al. Impulse control differentiates Internet gaming disorder from non-disordered but heavy Internet gaming use: Evidence from multiple behavioral and multimodal neuroimaging data. Computers in Human Behavior. 2022 May 1;130:107184. doi:10.1016/j.chb.2022.107184

22. Cyders MA, Smith GT. Emotion-based dispositions to rash action: Positive and negative urgency. Psychological Bulletin. 2008;134(6):807–28. doi:10.1037/a0013341

23. Sharma L, Markon KE, Clark LA. Toward a theory of distinct types of “impulsive” behaviors: A meta-analysis of self-report and behavioral measures. Psychological Bulletin. 2014;140(2):374–408. doi:10.1037/a0034418

24. Brand M, Wegmann E, Stark R, Müller A, Wölfling K, Robbins TW, et al. The Interaction of Person-Affect-Cognition-Execution (I-PACE) model for addictive behaviors: Update, generalization to addictive behaviors beyond internet-use disorders, and specification of the process character of addictive behaviors. Neuroscience and Biobehavioral Reviews. Elsevier Ltd; 2019. p. 1–10. doi:10.1016/j.neubiorev.2019.06.032 PubMed PMID: 31247240.

25. Warburton WA, Parkes S, Sweller N. Internet Gaming Disorder: Evidence for a Risk and Resilience Approach. International Journal of Environmental Research and Public Health. 2022 Jan;19(9):9. doi:10.3390/ijerph19095587

26. Ji Y, Yin MXC, Zhang AY, Wong DFK. Risk and protective factors of Internet gaming disorder among Chinese people: A meta-analysis. Aust N Z J Psychiatry. 2022 Apr;56(4):332–46. doi:10.1177/00048674211025703 PubMed PMID: 34250835.

27. Şalvarlı Şİ, Griffiths MD. The association between internet gaming disorder and impulsivity: A systematic review of literature. International Journal of Mental Health and Addiction. 2022;20(1):92–118. doi:10.1007/s11469-019-00126-w

28. Nuske J, Nuske L, Stevens MWR, Billieux J, Delfabbro PH, Hides L, et al. The association between gaming disorder and impulsivity: A systematic review and meta-analysis. Aust N Z J Psychiatry. 2025 Nov 12;00048674251388779. doi:10.1177/00048674251388779

29. Gao YX, Wang JY, Dong GH. The prevalence and possible risk factors of internet gaming disorder among adolescents and young adults: Systematic reviews and meta-analyses. Journal of Psychiatric Research. 2022 Oct 1;154:35–43. doi:10.1016/j.jpsychires.2022.06.049

30. Bäcklund C, Elbe P, Gavelin HM, Sörman DE, Ljungberg JK. Gaming motivations and gaming disorder symptoms: A systematic review and meta-analysis. Journal of Behavioral Addictions. 2022 Sep 12;11(3):667–88. doi:10.1556/2006.2022.00053

31. Odgers CL, Jensen M. Adolescent Mental Health in the Digital Age: Facts, Fears and Future Directions. J Child Psychol Psychiatry. 2020 Mar;61(3):336–48. doi:10.1111/jcpp.13190 PubMed PMID: 31951670; PubMed Central PMCID: PMC8221420.

32. Laconi S, Pirès S, Chabrol H. Internet gaming disorder, motives, game genres and psychopathology. Computers in Human Behavior. 2017 Oct 1;75:652–9. doi:10.1016/j.chb.2017.06.012

33. Pontes HM, Schivinski B, Kannen C, Montag C. The interplay between time spent gaming and disordered gaming: A large-scale world-wide study. Social Science & Medicine. 2022 Mar 1;296:114721. doi:10.1016/j.socscimed.2022.114721

34. Mihara S, Higuchi S. Cross-sectional and longitudinal epidemiological studies of Internet gaming disorder: A systematic review of the literature. Psychiatry and Clinical Neurosciences. 2017;71(7):425–44. doi:10.1111/pcn.12532

35. Wang HY, Cheng C. The Associations Between Gaming Motivation and Internet Gaming Disorder: Systematic Review and Meta-analysis. JMIR Mental Health. 2022 Feb 17;9(2):e23700. doi:10.2196/23700

36. Yen JY, Liu TL, Wang PW, Chen CS, Yen CF, Ko CH. Association between Internet gaming disorder and adult attention deficit and hyperactivity disorder and their correlates: Impulsivity and hostility. Addictive Behaviors. 2017 Jan 1;64:308– 13. doi:10.1016/j.addbeh.2016.04.024

37. King DL, Chamberlain SR, Carragher N, Billieux J, Stein D, Mueller K, et al. Screening and assessment tools for gaming disorder: A comprehensive systematic review. Clinical Psychology Review. 2020 Apr 1;77:101831. doi:10.1016/j.cpr.2020.101831

38. Pujol J, Fenoll R, Forns J, Harrison BJ, Martínez-Vilavella G, Macià D, et al. Video gaming in school children: How much is enough? Annals of Neurology. 2016 Sep;80(3):424–33. doi:10.1002/ana.24745 PubMed PMID: 27463843.

39. Przybylski AK, Weinstein N. A Large-Scale Test of the Goldilocks Hypothesis: Quantifying the Relations Between Digital-Screen Use and the Mental Well-Being of Adolescents. Psychol Sci. 2017 Feb 1;28(2):204–15. doi:10.1177/0956797616678438

40. Achab S, Nicolier M, Mauny F, Monnin J, Trojak B, Vandel P, et al. Massively multiplayer online role-playing games: comparing characteristics of addict vsnon-addict online recruited gamers in a French adult population. BMC Psychiatry. 2011 Dec;11(1):144. doi:10.1186/1471-244X-11-144

41. Vuorre M, Orben A, Przybylski AK. There Is No Evidence That Associations Between Adolescents’ Digital Technology Engagement and Mental Health Problems Have Increased. Clinical Psychological Science. 2021 Sep 1;9(5):823– 35. doi:10.1177/2167702621994549

42. Hussain Z, Griffiths MD. Excessive use of Massively Multi-Player Online Role-Playing Games: A Pilot Study. Int J Ment Health Addiction. 2009 Feb 20;7(4):4. doi:10.1007/s11469-009-9202-8

43. Laconi S, Pirès S, Chabrol H. Internet gaming disorder, motives, game genres and psychopathology. Computers in Human Behavior. 2017 Oct 1;75:652–9. doi:10.1016/j.chb.2017.06.012

44. Pontes HM, Király O, Demetrovics Z, Griffiths MD. The Conceptualisation and Measurement of DSM-5 Internet Gaming Disorder: The Development of the IGD-20 Test. PLOS ONE. 2014 Oct 14;9(10):10. doi:10.1371/journal.pone.0110137

45. Choo H, Gentile DA, Sim T, Li D, Khoo A, Liau AK. Pathological video-gaming among Singaporean youth. Ann Acad Med Singap. 2010 Nov;39(11):822–9. PubMed PMID: 21165520.

46. Paulus FW, Ohmann S, von Gontard A, Popow C. Internet gaming disorder in children and adolescents: a systematic review. Developmental Medicine & Child Neurology. 2018;60(7):645–59. doi:10.1111/dmcn.13754

47. Gentile DA, Choo H, Liau A, Sim T, Li D, Fung D, et al. Pathological video game use among youths: A two-year longitudinal study. Pediatrics. 2011 Feb;127(2). doi:10.1542/peds.2010-1353 PubMed PMID: 21242221.

48. Petry NM, Rehbein F, Gentile DA, Lemmens JS, Rumpf HJ, Mößle T, et al. An international consensus for assessing internet gaming disorder using the new DSM-5 approach. Addiction. 2014 Sep;109(9):1399–406. doi:10.1111/add.12457 PubMed PMID: 24456155.

49. Liau AK, Choo H, Li D, Gentile DA, Sim T, Khoo A. Pathological video-gaming among youth: A prospective study examining dynamic protective factors. Addiction Research & Theory. 2015;23(4):301–8. doi:10.3109/16066359.2014.987759

50. Petry NM, Rehbein F, Gentile DA, Lemmens JS, Rumpf HJ, Mößle T, et al. An international consensus for assessing internet gaming disorder using the new DSM-5 approach. Addiction. 2014;109(9):9. doi:10.1111/add.12457

51. Lemmens JS, Valkenburg PM, Gentile DA. The Internet Gaming Disorder Scale. Psychological Assessment. 2015 Jun;27(2):567–82. doi:10.1037/pas0000062

52. Király O, Sleczka P, Pontes HM, Urbán R, Griffiths MD, Demetrovics Z. Validation of the Ten-Item Internet Gaming Disorder Test (IGDT-10) and evaluation of the nine DSM-5 Internet Gaming Disorder criteria. Addict Behav. 2017 Jan;64:253–60. doi:10.1016/j.addbeh.2015.11.005 PubMed PMID: 26632194.

53. Király O, Bőthe B, Ramos-Diaz J, Rahimi-Movaghar A, Lukavska K, Hrabec O, et al. Ten-Item Internet Gaming Disorder Test (IGDT-10): Measurement invariance and cross-cultural validation across seven language-based samples. Psychology of Addictive Behaviors. 2019;33(1):1. doi:10.1037/adb0000433

54. Paschke K, Sack PM, Thomasius R. Validity and Psychometric Properties of the Internet Gaming Disorder Scale in Three Large Independent Samples of Children and Adolescents. Int J Environ Res Public Health. 2021 Feb;18(3):1095. doi:10.3390/ijerph18031095 PubMed PMID: 33530635; PubMed Central PMCID: PMC7908108.

55. King DL, Chamberlain SR, Carragher N, Billieux J, Stein D, Mueller K, et al. Screening and assessment tools for gaming disorder: A comprehensive systematic review. Clin Psychol Rev. 2020 Apr;77:101831. doi:10.1016/j.cpr.2020.101831 PubMed PMID: 32143109.

56. Hayes AF, Coutts JJ. Use Omega Rather than Cronbach’s Alpha for Estimating Reliability. But…. Communication Methods and Measures. 2020 Jan 2;14(1):1–24. doi:10.1080/19312458.2020.1718629

57. Dunn TJ, Baguley T, Brunsden V. From alpha to omega: a practical solution to the pervasive problem of internal consistency estimation. Br J Psychol. 2014 Aug;105(3):399–412. doi:10.1111/bjop.12046 PubMed PMID: 24844115.

58. Patton JH, Stanford MS, Barratt ES. Factor structure of the Barratt impulsiveness scale. J Clin Psychol. 1995 Nov;51(6):768–74. Located at: 8778124

59. Glezer H. Antecedents and correlates of marriage and family attitudes in young Australian men and women. In: Proceedings of the Twentieth International CFR Seminar on Social Change and Family Policies,. Melbourne.; 1984.

60. Simonsohn U, Simmons JP, Nelson LD. Specification curve analysis. Nat Hum Behav. 2020 Jul 27;4(11):1208–14. doi:10.1038/s41562-020-0912-z

61. Field A. Discovering statistics using IBM SPSS statistics: and sex and drugs and rock “n” roll. 4th edition. Los Angeles London New Delhi Singapore Washington DC: Sage; 2013. 915 p. (MobileStudy).

62. Thorell LB, Burén J, Wiman JS, Sandberg D, Nutley SB. Longitudinal associations between digital media use and ADHD symptoms in children and adolescents: a systematic literature review. European Child and Adolescent Psychiatry. Springer Science and Business Media Deutschland GmbH; 2024. p. 2503–26. doi:10.1007/s00787-022-02130-3 PubMed PMID: 36562860.

63. Swing EL, Gentile DA, Anderson CA, Walsh DA. Television and video game exposure and the development of attention problems. Pediatrics. 2010;126(2):214–21. doi:10.1542/peds.2009-1508

64. Gentile DA, Swing EL, Lim CG, Khoo A. Video game playing, attention problems, and impulsiveness: Evidence of bidirectional causality. Psychology of Popular Media Culture. 2012;1:62–70. doi:10.1037/a0026969

65. Tiraboschi GA, Fitzpatrick C, Kim HS, Superbia-Guimarães L, Kosak LA, Garon-Carrier G. Bidirectional associations between video game playing and ADHD symptoms among school-aged children. Computers in Human Behavior Reports. 2025 Aug 1;19:100740. doi:10.1016/j.chbr.2025.100740

66. Barlett C, Branch O, Rodeheffer C, Harris R. How long do the short-term violent video game effects last? Aggressive behavior. 2009;35:225–36. Located at: 19206102. doi:10.1002/ab.20301

67. Międzobrodzka E, Waiyaki F, Buczny J, Konijn E. Stop or Go? Playing Violent Games Reduces Inhibitory Control in Adolescents. 2021. doi:10.31234/osf.io/2anrt

68. Hummer T, Kronenberger W, Wang Y, Mathews V. Decreased Prefrontal Activity During a Cognitive Inhibition Task Following Violent Video Game Play: A Multi-Week Randomized Trial. Psychology of Popular Media Culture. 2017 May 11;8:63–75. doi:10.1037/ppm0000141

69. Anderson CA, Bushman BJ. Media Violence and the General Aggression Model. Journal of Social Issues. 2018 Jun;74(2):386–413. doi:10.1111/josi.12275

70. Ferguson CJ. Aggressive video games research emerges from its replication crisis (Sort of). Current Opinion in Psychology. 2020 Dec 1;Cyberpsychology36:1–6. doi:10.1016/j.copsyc.2020.01.002

71. Ferguson C, Copenhaver A, Markey P. Reexamining the Findings of the American Psychological Association’s 2015 Task Force on Violent Media: A Meta-Analysis. Perspectives on Psychological Science. 2020 Aug 10;15:174569162092766. doi:10.1177/1745691620927666

72. Kühn S, Kugler DT, Schmalen K, Weichenberger M, Witt C, Gallinat J. Does playing violent video games cause aggression? A longitudinal intervention study. Molecular Psychiatry. 2019 Aug;24(8):1220–34. doi:10.1038/s41380-018-0031-7 PubMed PMID: 29535447.

73. Moshel ML, Warburton WA, Batchelor J, Bennett JM, Ko KY. Neuropsychological Deficits in Disordered Screen Use Behaviours: A Systematic Review and Meta-analysis. Neuropsychol Rev. 2024;34(3):791–822. doi:10.1007/s11065-023-09612-4 PubMed PMID: 37695451; PubMed Central PMCID: PMC11473542.

74. Argyriou E, Davison CB, Lee TTC. Response inhibition and internet gaming disorder: A meta-analysis. Addict Behav. 2017;71:54–60. doi:10.1016/j.addbeh.2017.02.026

75. Ioannidis K, Hook R, Goudriaan AE, Vlies S, Fineberg NA, Grant JE, et al. Cognitive deficits in problematic internet use: meta-analysis of 40 studies. British Journal of Psychiatry. Cambridge University Press; 2019. p. 639–46. doi:10.1192/bjp.2019.3 PubMed PMID: 30784392.

76. Choi E, Shin SH, Ryu JK, Jung KI, Hyun Y, Kim J, et al. Association of extensive video gaming and cognitive function changes in brain-imaging studies of pro gamers and individuals with gaming disorder: Systematic literature review. JMIR Serious Games. JMIR Publications Inc.; 2021. doi:10.2196/25793

77. Deleuze J, Long J, Liu TQ, Maurage P, Billieux J. Passion or addiction? Correlates of healthy versus problematic use of videogames in a sample of French-speaking regular players. Addictive Behaviors. 2018 Jul;82:114–21. doi:10.1016/j.addbeh.2018.02.031

78. Billieux J, Flayelle M, Rumpf HJ, Stein DJ. High Involvement Versus Pathological Involvement in Video Games: a Crucial Distinction for Ensuring the Validity and Utility of Gaming Disorder. Curr Addict Rep. 2019 Sep 1;6(3):323–30. doi:10.1007/s40429-019-00259-x

79. Charlton JP, Danforth IDW. Distinguishing addiction and high engagement in the context of online game playing. Computers in Human Behavior. 2007 May 1;Including the Special Issue: Avoiding Simplicity, Confronting Complexity: Advances in Designing Powerful Electronic Learning Environments23(3):1531–48. doi:10.1016/j.chb.2005.07.002

80. Twenge JM, Martin GN, Campbell WK. Decreases in psychological well-being among American adolescents after 2012 and links to screen time during the rise of smartphone technology. Emotion. 2018;18(6):765–80. doi:10.1037/emo0000403

81. Sallis JF, Prochaska JJ, Taylor WC. A review of correlates of physical activity of children and adolescents. Med Sci Sports Exerc. 2000 May;32(5):963–75. doi:10.1097/00005768-200005000-00014 PubMed PMID: 10795788.

82. Yu L, Shek DTL. Internet addiction in Hong Kong adolescents: a three-year longitudinal study. J Pediatr Adolesc Gynecol. 2013 Jun;26(3 Suppl):S10–17. doi:10.1016/j.jpag.2013.03.010 PubMed PMID: 23683821.

83. Yu C, Li X, Zhang W. Predicting adolescent problematic online game use from teacher autonomy support, basic psychological needs satisfaction, and school engagement: a 2-year longitudinal study. Cyberpsychol Behav Soc Netw. 2015 Apr;18(4):228–33. doi:10.1089/cyber.2014.0385 PubMed PMID: 25803769.

84. Zhu J, Zhang W, Yu C, Bao Z. Early adolescent Internet game addiction in context: How parents, school, and peers impact youth. Computers in Human Behavior. 2015;50:159–68. doi:10.1016/j.chb.2015.03.079

85. Sugaya N, Shirasaka T, Takahashi K, Kanda H. Bio-psychosocial factors of children and adolescents with internet gaming disorder: A systematic review. BioPsychoSocial Medicine. BioMed Central Ltd.; 2019. doi:10.1186/s13030-019-0144-5

86. King DL, Chamberlain SR, Carragher N, Billieux J, Stein D, Mueller K, et al. Screening and assessment tools for gaming disorder: A comprehensive systematic review. Clinical Psychology Review. 2020 Apr 1;77:101831. doi:10.1016/j.cpr.2020.101831

87. Stevens MWR, Radünz M, Galanis C, Quinney B, Zajac I, Billieux J, et al. Red Box, Green Box: Psychometric evaluation of a self-report behavioral frequency measurement approach for behavioral addictions research. Addiction. 2026;121(2):429–39. doi:10.1111/add.70192

88. Park JJ, Booth N, Bagot KL, Rodda SN. A brief internet-delivered intervention for the reduction of gaming-related harm: A feasibility study. Computers in Human Behavior Reports. 2020 Aug 1;2:100027. doi:10.1016/j.chbr.2020.100027

89. Reangsing C, Wongchan W, Trakooltorwong P, Thaibandit J, Oerther S. Effects of cognitive behavioral therapy (CBT) on addictive symptoms in individuals with internet gaming disorders: A systematic review and meta-analysis. Psychiatry Research. 2025 Jun 1;348:116425. doi:10.1016/j.psychres.2025.116425

90. Harpas I, Stevens M, Radunz M, Williamson P, Hamamura T, Svendsen O, et al. Treatment of gaming disorder: A systematic review and meta-analysis. Psychiatry Research. 2025 Dec 1;354:116783. doi:10.1016/j.psychres.2025.116783

91. Stevens MWR, King DL, Dorstyn D, Delfabbro PH. Cognitive-behavioral therapy for Internet gaming disorder: A systematic review and meta-analysis. Clin Psychol Psychother. 2019 Mar;26(2):191–203. doi:10.1002/cpp.2341 PubMed PMID: 30341981.

92. Bediou B, Rich M, Bavelier D. Digital media and cognitive development [Internet]. 2020 Oct 15. 10.1787/3b071e13-en

